# Disease profile and plasma neutralizing activity of post-vaccination Omicron BA.1 infection in Tianjin, China: a retrospective study

**DOI:** 10.1101/2022.04.09.22273653

**Authors:** Hong Zheng, Yunlong Cao, Xiaosu Chen, Fengmei Wang, Ye Hu, Weiliang Song, Yangyang Chai, Qingqing Gu, Yansong Shi, Yingmei Feng, Shuxun Liu, Yan Xie, Xiaoliang Sunney Xie, Wentao Jiang, Zhongyang Shen

## Abstract

**Background:** SARS-CoV-2 Omicron variant BA.1 first emerged on the Chinese mainland in January 2022 in Tianjin and caused a large wave of infections. During mass PCR testing, a total of 430 cases infected with Omicron were recorded between January 8 and February 7, 2022, with no new infections detected for the following 16 days. Most patients had been vaccinated with SARS-CoV-2 inactivated vaccines. The disease profile associated with BA.1 infection, especially after vaccination with inactivated vaccines, is unclear. Whether BA.1 breakthrough infection after receiving inactivated vaccine could create a strong enough humoral immunity barrier against Omicron is not yet investigated.

**Methods:** We collected the clinical information and vaccination history of the 430 COVID-19 patients infected with Omicron BA.1. Re-positive cases and inflammation markers were monitored during the patient’s convalescence phase. Ordered multiclass logistic regression model was used to identify risk factors for COVID-19 disease severity. Authentic virus neutralization assays against SARS-CoV-2 wildtype, Beta and Omicron BA.1 were conducted to examine the plasma neutralizing titers induced after post-vaccination Omicron BA.1 infection, and were compared to a group of uninfected healthy individuals who were selected to have a matched vaccination profile.

**Findings:** Among the 430 patients, 316 (73.5%) were adults with a median age of 47 years, and 114 (26.5%) were under-age with a median age of 10 years. Female and male patients account for 55.6% and 44.4%, respectively. Most of the patients presented with mild (47.7%) to moderate diseases (50.2%), with only 2 severe cases (0.5%) and 7 (1.6%) asymptomatic infections. No death was recorded. 341 (79.3%) of the 430 patients received inactivated vaccines (54.3% BBIBP-CorV vs. 45.5% CoronaVac), 49 (11.4%) received adenovirus-vectored vaccines (Ad5-nCoV), 2 (0.5%) received recombinant protein subunit vaccines (ZF2001), and 38 (8.8%) received no vaccination. No vaccination is associated with a substantially higher ICU admission rate among Omicron BA.1 infected patients (2.0% for vaccinated patients vs. 23.7% for unvaccinated patients, P<0.001). Compared with adults, child patients presented with less severe illness (82.5% mild cases for children vs. 35.1% for adults, P<0.001), no ICU admission, fewer comorbidities (3.5% vs. 53.2%, P<0.001), and less chance of turning re-positive on nucleic acid tests (12.3% vs. 22.5%, P=0.019). For adult patients, compared with no prior vaccination, receiving 3 doses of inactivated vaccine was associated with significantly lower risk of severe disease (OR 0.227 [0.065-0.787], P=0.020), less ICU admission (OR 0.023 [0.002-0.214], P=0.001), lower re-positive rate on PCR (OR 0.240 [0.098-0.587], P=0.002), and shorter duration of hospitalization and recovery (OR 0.233 [0.091-0.596], P=0.002). At the beginning of the convalescence phase, patients who had received 3 doses of inactivated vaccine had substantially lower systemic immune-inflammation index (SII) and C-reactive protein than unvaccinated patients, while CD4+/CD8+ ratio, activated Treg cells and Th1/Th2 ratio were higher compared to their 2-dose counterparts, suggesting that receipt of 3 doses of inactivated vaccine could step up inflammation resolution after infection. Plasma neutralization titers against Omicron, Beta, and wildtype significantly increased after breakthrough infection with Omicron. Moderate symptoms were associated with higher plasma neutralization titers than mild symptoms. However, vaccination profiles prior to infection, whether 2 doses versus 3 doses or types of vaccines, had no significant effect on post-infection neutralization titer. Among recipients of 3 doses of CoronaVac, infection with Omicron BA.1 largely increased neutralization titers against Omicron BA.1 (8.7x), Beta (4.5x), and wildtype (2.2x), compared with uninfected healthy individuals who have a matched vaccination profile.

**Interpretation:** Receipt of 3-dose inactivated vaccines can substantially reduce the disease severity of Omicron BA.1 infection, with most vaccinated patients presenting with mild to moderate illness. Child patients present with less severe disease than adult patients after infection. Omicron BA.1 convalescents who had received inactivated vaccines showed significantly increased plasma neutralizing antibody titers against Omicron BA.1, Beta, and wildtype SARS-CoV-2 compared with vaccinated healthy individuals.

**Funding:** This research is supported by Changping Laboratory (CPL-1233) and the Emergency Key Program of Guangzhou Laboratory (EKPG21-30-3), sponsored by the Ministry of Science and Technology of the People’s Republic of China.

**Research in context:** *Evidence before this study:* Previous studies (many of which have not been peer-reviewed) have reported inconsistent findings regarding the effect of inactivated vaccines against the Omicron variant. On Mar 6, 2022, we searched PubMed with the query “(SARS-CoV-2) AND ((Neutralisation) OR (Neutralisation)) AND ((Omicron) OR (BA.1)) AND (inactivated vaccine)”, without date or language restrictions. This search identified 18 articles, of which 13 were directly relevant. Notably, the participants in many of these studies have received only one or two doses of inactivated vaccine with heterologous booster vaccination; other studies have a limited number of participants receiving inactivated vaccines.

*Added value of this study:* To date, this is the first study to report on the protective effect of inactivated vaccines against the severe disease caused by the Omicron variant. We examine and compare the disease profile of adults and children. Furthermore, we estimate the effect of post-vaccination omicron infection on plasma neutralization titers against Omicron and other SARS-COV-2 variants. Specifically, the disease profile of Omicron convalescents who had received two-dose primary series of inactivated vaccines with or without a booster dose prior to infection is compared with unvaccinated patients. We also analyzed the effect of infection on neutralizing activity by comparing vaccinated convalescents with vaccinated healthy individuals with matched vaccination profiles.

*Implications of all the available evidence:* Compared with adults, child patients infected with Omicron tend to present with less severe disease and are less likely to turn re-positive on nucleic acid tests. Receipt of two-dose primary series or three doses of inactivated vaccine is a protective factor against severe disease, ICU admission, re-positive PCR and longer hospitalization. The protection afforded by a booster dose is stronger than two-dose primary series alone. Besides vaccination, infection with Omicron is also a key factor for elevated neutralizing antibody titers, enabling cross-neutralization against Omicron, wildtype (WT) and the Beta variant.

## Introduction

Coronavirus disease 2019 (COVID-19), an acute respiratory infectious disease caused by SARS-CoV-2, hit the world by surprise over two years ago and still is wreaking havoc worldwide. Multiple variants of concern (VOCs) have emerged with varied transmissibility and immune evasiveness, capable of causing breakthrough infections in vaccinated individuals and those with prior infection. Currently, the highly mutated Omicron is the dominant variant circulating in multiple countries and regions, and is still on the rise,^1, 2^ Omicron carries over 30 mutations on its spike (S) protein, including 15 mutations on the receptor-binding domain (RBD) compared with 2 mutations for the Delta variant.^3^ Several mutations harbored by Omicron are shared with the Delta and Alpha variant, which is closely related to its increased transmissibility and immune evasiveness.

On January 8, 2022, the first non-imported Omicron case (BA.1) on the Chinese mainland was reported in Tianjin. Four rounds of mass testing of its approximately 14 million residents were launched on January 9, 12, 15 and 20, respectively, followed by multiple targeted testing in high-risk areas. As of February 7, 2022, a total of 430 individuals were tested positive for Omicron BA.1, characterized by family clusters (263/430, 61.16%) and community transmission (93/430, 21.63%). Early in the outbreak, cluster cases among children aged between 7-12 years were reported in childcare centers and schools.

Mass vaccination programs have been rolled out globally over the past 2 years. In the Chinese mainland, the most delivered vaccines are inactivated vaccines. As of January 8, 2022, when Omicron first emerged in Tianjin, up to 93.2% of its residents had been vaccinated to a varied extent.^4^ However, the effect of inactivated vaccines on infection and disease severity remains unclear. We investigated the protection conferred by vaccination against Omicron by examining the breakthrough infections among recipients of vaccines as compared with unvaccinated cases. Our study population comprised 430 cases in Tianjin during the outbreak of Omicron BA.1 infection in January, of which 79.3% were inoculated with inactivated vaccines, namely CoronaVac or BBIBP-CorV.

## Methods

### Data Sources

This study analyzed clinical and demographic data on the 430 Omicron patients reported by Tianjin Municipal Health Commission between January 8 and February 7, 2022, including COVID-19 vaccination history, laboratory tests, clinical symptoms, ICU admission and death. These data were extracted from the medical records obtained from Tianjin Haihe Hospital and Tianjin First Central Hospital where the patients received treatment after diagnosis.

### Laboratory Confirmation

In this study, laboratory diagnosis was done in two designated COVID-19 hospitals: Tianjin Haihe Hospital and Tianjin First Central Hospital. Nucleic acid was extracted from respiratory samples using commercial kits (Zybio, 5203050). Reverse Transcription-Polymerase Chain Reaction (RT-PCR) was performed following the WHO protocol to detect two target genes, the open reading frame of 1ab (ORF1ab) and the nucleocapsid protein (N).^5^ ORF1ab forward primer (F): CCCTGTGGGTTTTACACTTAA, reverse primer (R): ACGATTGTGCATCAGCTGA, probe (P): 5’-FAM-CCGTCTGCGGTATGTGGAAAGGTTATGG-BHQ1-3’. N forward primer: GGGGAACTTCTCCTGCTAGAAT, reverse primer: CAGACATTTTGCTCTCAAGCTG, probe: 5’-FAM-TTGCTGCTGCTTGACAGATT-TAMRA-3’. 40 cycles of amplification (50°C for 10 min, 90°C for 5 min, 95°C for 10 s; 55°C for 40 s) were performed. A cycle threshold value (Ct-value) less than 37 with an S-shape amplification curve was defined as positive, and a Ct-value no less than 40 without a S-shape amplification curve was defined as negative. In the case of 37 ≤ Ct < 40, retesting would be recommended. If both target genes (ORF1ab and N) are tested positive on real-time RT-PCR for the same sample, the result would be regarded as positive; if only one of the two target genes is tested positive, resampling and retesting would be required. If one of the two target genes is tested positive for two samples, the result would be positive.

### Plasma Isolation

Peripheral blood was collected from 140 Omicron BA.1 convalescent patients 28 days after discharge and 114 healthy recipients of 3 doses of inactivated vaccines (CoronaVac) around 90 days after the booster shot. Whole blood samples were then diluted with PBS+2%FBS at 1:1 for Ficoll gradient centrifugation (Cytiva, 17-1440-03). Plasma was collected from the supernatants and restored at −80°C.

### Authentic neutralization assay

A neutralization assay of authentic SARS-CoV-2 was performed using a cytopathic effect (CPE) assay. Briefly, various concentrations (2-fold serial dilution using DMEM) of plasma were mixed with the same volume of 100 PFU of authentic SARS-CoV-2 and incubated at 37°C for 1 h. The mixture was added to a monolayer of Vero-E6 cells (5×10^3^ cells per well) in a 96-well plate and incubated for 1 h at 37°C. The supernatant was removed, and 200 μL of DMEM supplied with 2% (v/v) FBS was added to the infected cells. After incubation at 37°C supplied with 5% CO2 for 5 days, all wells were examined for the CPE effect. All experiments were performed in a Biosafety Level 3 facility. Neutralization titers were calculated by the Spearman-Karber method.

### Outcomes

The primary endpoints were COVID-19 severity and neutralizing antibody titers. COVID-19 disease severity was defined as asymptomatic, mild, moderate, severe and critical according to WHO living guidance for clinical management of COVID-19.^6^

Secondary endpoints were intensive care unit (ICU) admission, re-positive results on nucleic acid tests, and duration of hospitalization and recovery. PCR re-positive was defined as PCR Ct value < 40 after two independent PCR-negative results at an interval of more than 24 hours. Duration of hospitalization and recovery was defined as the days spent in Tianjin Haihe Hospital and Tianjin First Central Hospital, respectively. All patients, including asymptomatic and mild cases, were hospitalized in Tianjin Haihe Hospital upon positive PCR results. Patients were discharged from Tianjin Haihe Hospital if the following criteria were met: 1) body temperature restored and stayed normal for over 3 days; 2) respiratory symptoms significantly relieved; 3) acute exudation substantially resolved on imaging study of the lungs; 4) negative on two consecutive PCR tests (at an interval of at least 24 hours) of samples collected from the respiratory tract. For patients whose PCR assays remained positive for over 4 weeks after criteria 1), 2) and 3) had been met, antibody assay and virus culture were applied to assess the risk of transmission before deciding whether these patients could be discharged.

Discharged patients from Tianjin Haihe Hospital were then transferred to Tianjin First Central Hospital for at least 14 days under medical observation. Patients received PCR assays on the 1^st^, 7^th^ and 14^th^ days after being transferred to Tianjin First Central Hospital. After 14 days of observation, patients with negative results on PCR and without other conditions in need of hospitalization were discharged. Re-positive cases were required to yield negative on consecutive PCR assays at an interval of at least 24 hours.

### Study Oversight

This study was approved by the Tianjin Municipal Health Commission and the Ethics Committee of Tianjin First Central Hospital (Ethics committee archiving No. 2022N045KY). All patients/participants provided their written informed consent to have their clinical information collected for study purposes and the data generated from the study published. All the authors contributed to data collection and analysis, discussion and interpretation of the results. All the authors read and approved the final manuscript.

### Statistical Analysis

Continuous variables were shown in medians and interquartile ranges (IQR), and the Mann-Whitney U test was used to analyze the differences between the two groups. Categorical variables were summarized as counts and percentages, and analyzed by Pearson’s χ2 test. Ordered multi-class Logistic regression model was used to analyze the relations between age/gender/receipt of inactivated vaccine and COVID-19 severity. All the analyses were conducted using the SPSS software, version 22.0. Two-sided P<0.05 was used in tests of significance.

## Results

### Demographic and Clinical Characteristics

During citywide mass testing, a total of 430 cases infected with Omicron BA.1 were recorded between January 8 and February 7, 2022, with no new infections detected for the following 16 days, resulting in a period prevalence of 3.10 cases per 100,000 people. The demographic and clinical characteristics of the patients are summarized in Table 1.

**Table 1.**
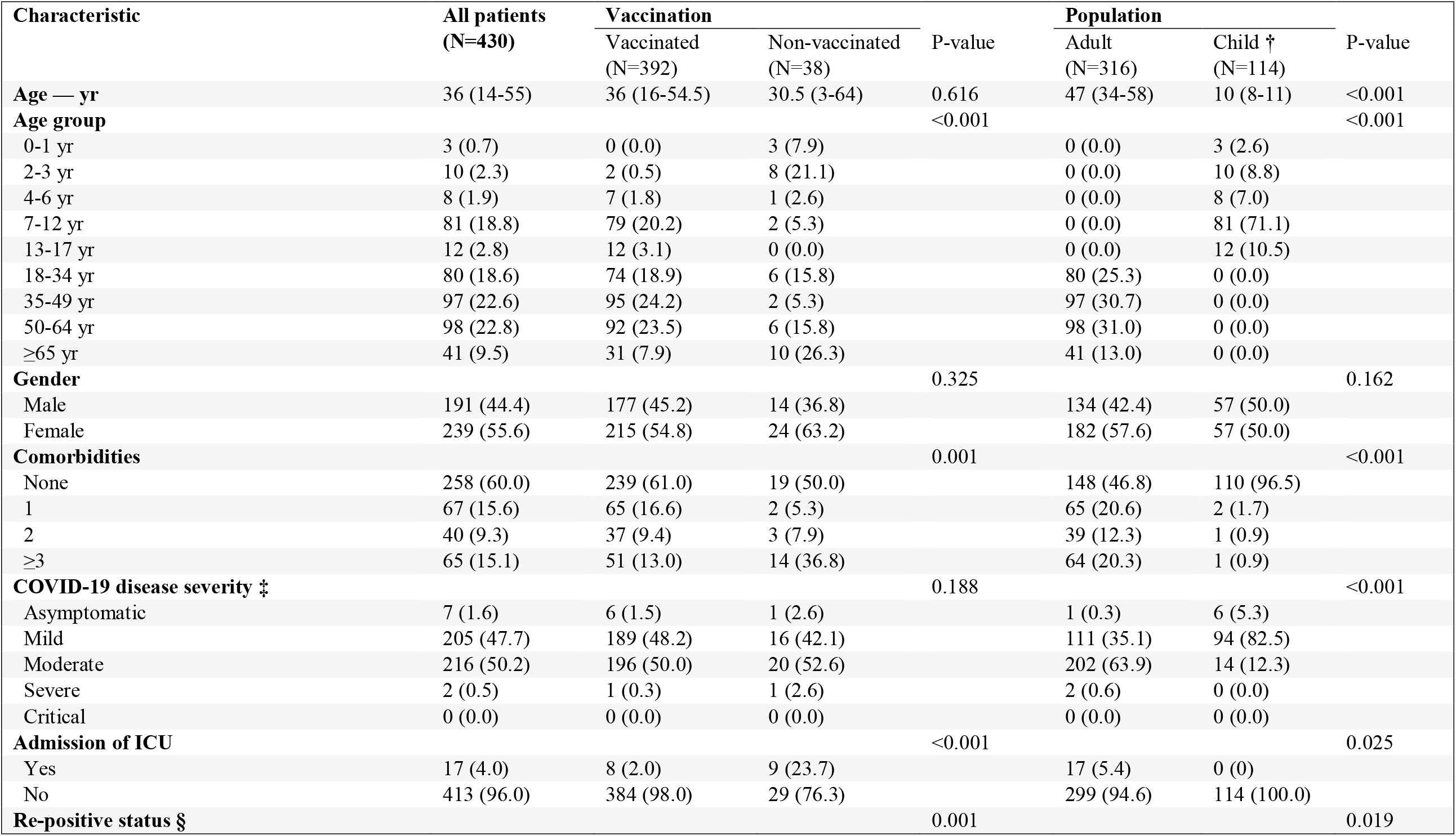
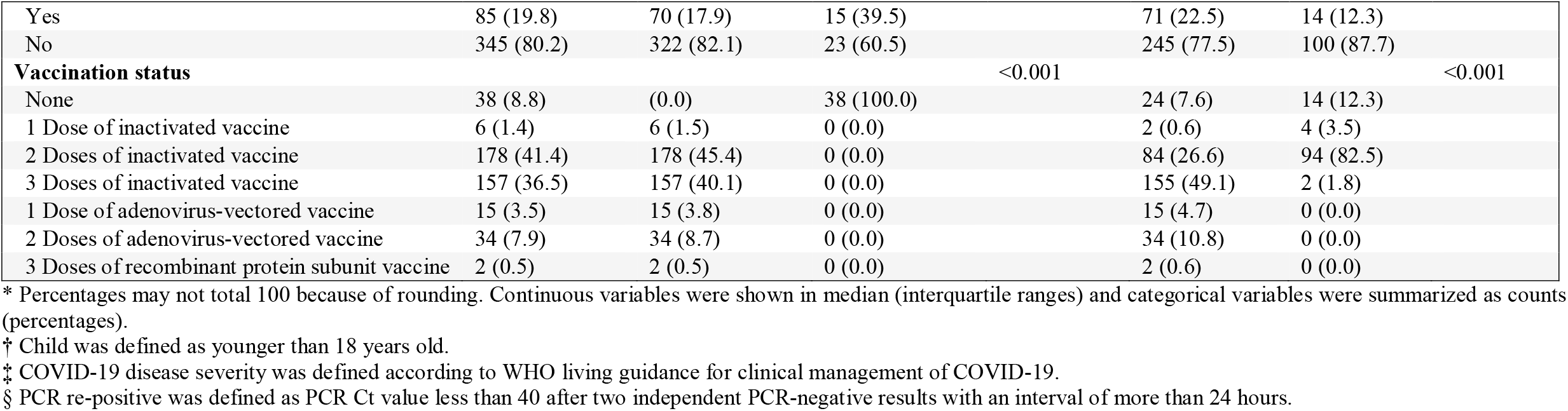
Characteristics of Omicron infected patients *.

The median age of the 430 patients was 36 years (interquartile range [IQR],14-55), with 26.5% under 18. Female and male patients account for 55.6% and 44.4%, respectively. The most common onset symptoms were cough (37.0%) and fever (30.2%); parageusia (1.4%), heterosmia (0.9%), diarrhea (0.9%) and rash (0.2%) were rare (Table S1). 40.0% of the patients had at least one comorbidity, with hypertension (17.0%) and abnormal liver function (16.0%) being the most common (Table S2). Patients with a history of SARS-COV-2 vaccination and child patients had fewer comorbidities as compared with vaccination-naïve patients (39.0% vs. 50.0%, P=0.001) and adult patients (3.5% vs. 53.2%, P<0.001), respectively.

Most patients presented with mild (47.7%) to moderate (50.2%) illness, with only 2 (0.5%) severe cases and no critical cases. A vast majority (82.5%) of the child patients had mild symptoms, with 12.3% moderate, 5.3% asymptomatic and no severe or critical cases. Adult patients were predominantly moderate cases (63.9%), followed by mild (35.1%), while severe and asymptomatic cases only accounted for 0.6% and 0.3%, respectively. Patients without comorbidities manifested less severe symptoms compared to those who had preexisting conditions at the time of Omicron infection (Table S3). 4.0% of the patients were admitted into ICU during hospitalization and were all adults. Vaccinated patients showed a substantially lower risk of ICU admission as compared with those unvaccinated (2.0% vs. 23.7%, P<0.001). After turning negative on nucleic acid tests, a significantly higher proportion of unvaccinated patients experienced re-positive results than vaccinated patients (39.5% vs. 17.9%, P=0.001). The re-positive rate was lower for child patients than adult patients (12.3% vs. 22.5%, P=0.019). Patients demonstrating continuous clinical symptoms during recovery had higher re-positive rates than those without (33.3% vs. 16.4%, P<0.001, Table S4).

### Effectiveness of inactivated vaccine against Omicron Variant

Of the 430 patients infected with Omicron BA.1, 341 (79.3%) received inactivated vaccines (54.3% BBIBP-CorV, 45.5% CoronaVac and 0.3% other), 49 (11.4%) received adenovirus-vectored vaccines (Ad5-nCoV), 2 (0.5%) recombinant protein subunit vaccine (ZF2001), and 38 (8.8%) were unvaccinated. The prevalence of infection among vaccinated individuals was 3.03/100,000, and 4.04/100,000 for the unvaccinated population (P=0.091). Given that Tianjin launched the vaccination programs for children aged between 3-11 years old since October 30, 2021, most of the infected children (82.5%) had received only 2 doses of inactivated vaccines, 3.5% received 1 dose, and 12.3% had received no vaccination.

Based on data from the 341 patients who had received inactivated vaccines and 38 patients who had no SARS-CoV-2 vaccination, we analyzed the correlation between the number of inactivated vaccine doses and disease severity (Table 2). It was found that when all age groups are considered together, 2 doses of inactivated vaccines associate with a larger proportion of asymptomatic to mild disease (less severity) compared with 3 doses (P=0.003), which may be because most under-age patients had received 2 doses of inactivated vaccine and presented with mild illness. In adult patients, 3 doses reduced disease severity compared with no vaccination (P=0.004). ICU admission rate was only 0.6% for patients who had received a booster dose of inactivated vaccine and 4.8% for those who only received two-dose primary series, both significantly lower than the 27.5% for unvaccinated patients (P<0.001).

**Table 2.**
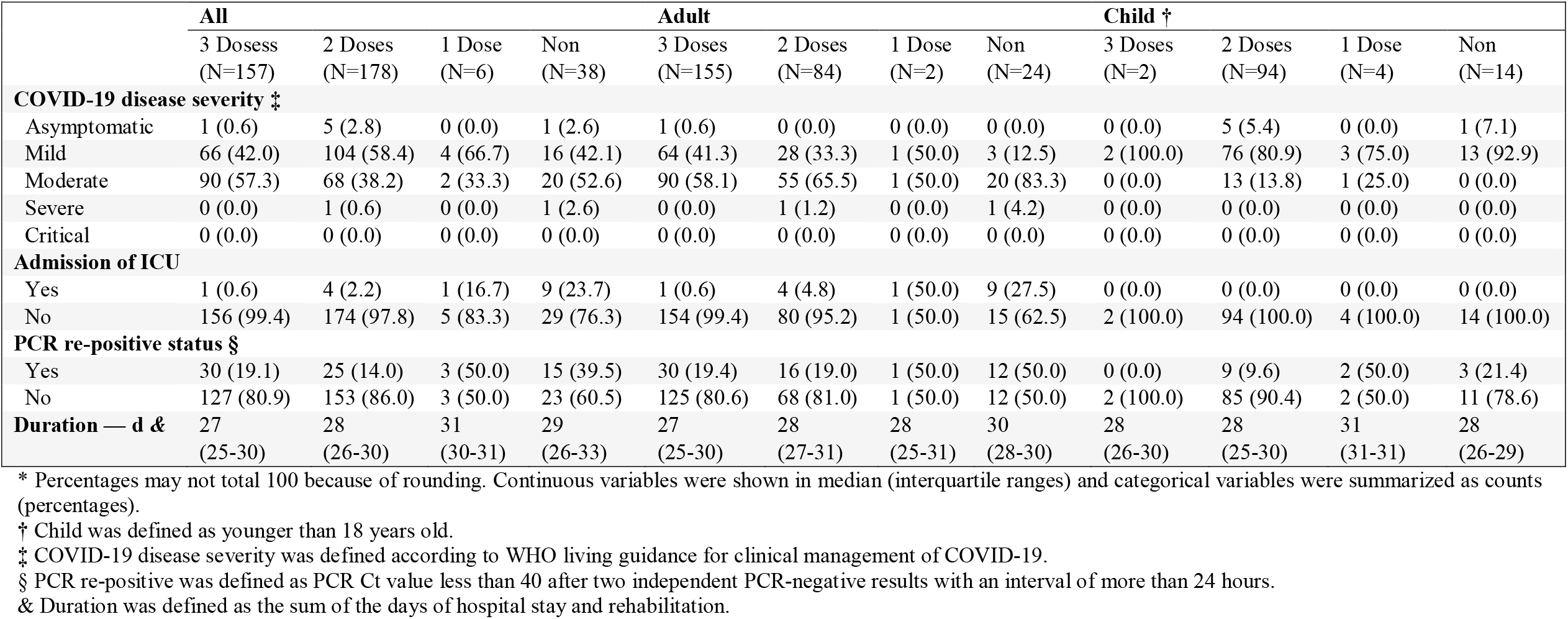
Correlation between inactivated vaccine doses and COVID-19 disease severity and progression *.

Receipt of two-dose primary series or three doses was associated with significantly lower rates of re-positive on nucleic acid tests. For patients across all age groups, the re-positive rate was 19.1% for those who had received 3 and 14.0% for those who had received 2 doses, both significantly lower than that of unvaccinated patients, which was 39.5% (P=0.008, P<0.001, respectively). For adult patients alone, receipt of 3 doses (19.4% vs. 50.0%, P=0.001) or 2 doses (19.0% vs. 50.0%, P=0.002) is also associated with lower re-positive rates than no vaccination. The same relations were not statistically significant in child patients, due to the limited sample size.

Patients who had received a booster dose of inactivated vaccine experienced a shorter period of hospitalization and recovery. For patients across all age groups, the duration of hospitalization and recovery was 2 days shorter for those who had received 3 doses of inactivated vaccine than for unvaccinated patients (P=0.009). For adult patients specifically, the duration of hospitalization and recovery for those who had received 3 doses (27 vs. 30 days, P=0.001) or 2 doses (28 vs. 30 days, P=0.026) of inactivated vaccine was shorter than unvaccinated patients; receipt of a booster dose was associated with fewer days of hospitalization and recovery than two-dose primary series alone (27 vs. 28, P=0.017).

Vaccination status also correlated with immunity and inflammation-related laboratory findings (Table 3). Compared with patients who had not been vaccinated against SARS-CoV-2, patients who had received 3 doses of inactivated vaccines showed significantly lower levels of the systemic immune-inflammatory index (SII) and C-reactive protein during the early stage of recovery after nucleic acid tests turned negative, suggesting that receipt of inactivated vaccine can step up inflammation resolution. Due to relatively lower levels of lymphocytes, neutrophil/lymphocyte ratio (NLR), platelet/lymphocyte ratio (PLR) and monocyte/lymphocyte ratio (MLR) was higher in patients inoculated with three doses of inactivated vaccine than that of patients vaccinated with two doses. T cell clustering indicated that the booster dose of inactivated vaccine led to a significant elevation of the CD4+/CD8+ ratio, the ratio of activated Treg cells and the Th1/Th2 ratio. Tests of liver and kidney functions suggest that the number of inactivated vaccine doses did not affect liver and kidney function (Table S5).

**Table 3.** Laboratory Findings *.

| Table 3. Laboratory Findings * |  |  |  |  |  |  |
| --- | --- | --- | --- | --- | --- | --- |
|  | 3 Doses | 2 Doses | 1 Dose | Non | P-value † |  |
| White-cell count — per mm<sup>3</sup> | 6320 (5490-7230) | 6310 (5300-7510) | 7995 (7720-10680) | 7420 (5130-8680) | 0.016 | 3 V.S. 1: 0.004<br>2 V.S. 1: 0.009 |
| Lymphocyte count — per mm<sup>3</sup> | 2080 (1650-2430) | 2350 (1910-2900) | 2930 (2640-3340) | 2600 (1530-4090) | 0.001 | 3 V.S.2: <0.001<br>3 V.S.1: 0.031 |
| Neutrophil count — per mm<sup>3</sup> | 3590 (2900-4230) | 3260 (2440-4050) | 4390 (3070-5720) | 3590 (2510-4690) | 0.045 | 3 V.S.2: 0.011 |
| Monocyte count — per mm<sup>3</sup> | 410 (350-500) | 410 (340-520) | 530 (390-590) | 490 (420-580) | 0.021 | 3 V.S. Non: 0.002<br>2 V.S. Non: 0.013 |
| Platelet count — per mm<sup>3</sup> | 262,000<br>(227,000-307,000) | 288,000<br>(238,000-326,000) | 316,000<br>(306,000-329,000) | 290,000<br>(237,000-386,000) | 0.008 | 3 V.S.2: 0.010<br>3 V.S.1: 0.011 |
| SII | 372 (239-642) | 492 (314-578) | 390 (248-550) | 468 (330-625) | 0.032 | 3 V.S.2: 0.003 |
| NLR | 1.72 (1.41-2.11) | 1.40 (1.00-1.96) | 1.40 (1.03-1.72) | 1.52 (0.93-2.36) | <0.001 | 3 V.S.2: <0.001 |
| PLR | 131 (104-162) | 114 (96-141) | 102 (97-118) | 107 (84-159) | 0.031 | 3 V.S.2: 0.009 |
| MLR | 0.20 (0.17-0.24) | 0.17 (0.14-0.22) | 0.17 (0.12-0.23) | 0.21 (0.14-0.30) | 0.003 | 3 V.S.2: <0.001 |
| C-reactive protein ≥10 mg/liter |  |  |  |  | <0.001 | 3 V.S. Non: <0.001<br>2 V.S. Non: 0.007 |
| Yes | 2 (1.3) | 6 (3.4) | 1 (16.7) | 6 (16.2) |  |  |
| No | 155 (98.7) | 171 (96.6) | 5 (83.3) | 31 (83.8) |  |  |
| Interleukin 6 ≥1.50 pg/milliliter |  |  |  |  | 0.309 |  |
| Yes | 45 (28.7) | 54 (30.5) | 1 (16.7) | 16 (43.2) |  |  |
| No | 112 (71.3) | 123 (69.5) | 5 (69.5) | 21 (56.8) |  |  |
| CD4<sup>+</sup> T cell count — per microliter | 0.71(0.44-1.08) | 0.75(0.47-1.15) | 0.78(0.61-1.01) | 0.65(0.39-1.16) | 0.841 |  |
| CD8<sup>+</sup> T cell count — per microliter | 0.77(0.48-1.30) | 0.80(0.51-1.45) | 0.48(0.34-3.32) | 0.97(0.48-2.30) | 0.508 |  |
| CD4<sup>+</sup>/CD8<sup>+</sup> | 1.68(1.27-2.14) | 1.47(1.17-1.78) | 1.67(1.47-1.99) | 1.56(1.35-2.29) | 0.034 | 3 V.S.2: 0.034 |
| Treg cell count — per microliter | 3.41(2.65-4.32) | 3.54(2.67-4.60) | 4.10(2.79-6.98) | 2.69(1.93-3.36) | 0.116 |  |
| Activated Treg cell count<br>— per microliter | 1.22(0.76-1.67) | 0.96(0.59-1.35) | 1.82(0.75-2.71) | 0.74(0.39-1.03) | 0.003 | 3 V.S. 2: 0.044<br>3 V.S. Non: 0.035 |
| Th1 cell count — per microliter | 25.59(21.32-31.84) | 22.17(17.96-27.86) | 23.72(16.80-30.07) | 28.23(21.05-33.22) | 0.001 | 3 V.S. 2: 0.001 |
| Th2 cell count — per microliter | 49.23(39.66-56.55) | 60.58(49.10-67.72) | 50.01(20.44-68.58) | 45.81(38.87-60.84) | <0.001 | 3 V.S. 2: <0.001 |
| Th1/Th2 | 0.55(0.38-0.79) | 0.37(0.28-0.55) | 0.59(0.24-1.60) | 0.62(0.36-0.76) | <0.001 | 3 V.S. 2: <0.001 |

\* Data regarding white-cell, lymphocyte, neutrophil, monocyte, platelet count, SII, NLR, PLR, MLR, and the measurement of C-reactive protein and Interleukin 6 were missing for 2 patients (0.5%). Data were missing for the proportion of T cell subsets in 106 patients (28.0%). SII was defined as follows: $SII = P \times N/L$ , where P, N and L are the preoperative peripheral blood platelet, neutrophil and lymphocyte counts per liter, respectively. NLR was defined as the absolute neutrophil count divided by the absolute lymphocyte count. PLR was defined as the platelet count divided by the number of lymphocytes. MLR was defined as the ratio of absolute monocyte count to absolute lymphocyte count. Continuous variables were shown in median (interquartile ranges) and categorical variables were summarized as counts (percentages).
† Only statistically significant results are shown here.

### Neutralization of the SARS-CoV-2 Omicron BA.1

We obtained plasma samples from 135 Omicron convalescent patients, 80 (59.3%) vaccinated with 3 doses of inactivated vaccine, 39 (28.9%) with 2 doses of inactivated vaccine, and 16 (11.8%) with 2 doses of Ad5-nCoV (adenovirus-vectored vaccine). Those who had received inactivated vaccines were vaccinated with CoronaVac (42 with three doses, 23 with two doses) or BBIBP-CorV (38 with three doses, 16 with two doses). The plasma samples were sampled one-month after hospitalization discharge.

We used authentic SARS-CoV-2 virus neutralization assays (CPE) to determine the plasma neutralizing antibody titers against WT, Beta and Omicron BA.1. For Omicron convalescent patients who had received 3 doses of inactivated vaccine, the geometric mean of 50% neutralizing titer (NT50) against WT was 4.2 and 8.4 times higher than the NT50 against Beta and Omicron, respectively (Figure 1a). Patients who had received 2 doses of inactivated vaccines showed a similar level of neutralizing antibody titers to those vaccinated with 3 doses (Figure 1a). Patients who had received inactivated vaccine, BBIBP-CorV or CoronaVac, displayed a similar level of neutralizing antibody titers. Those who had received the adenovirus-vectored vaccine (Ad5-nCoV) showed higher NT50 against WT, Beta and Omicron BA.1 compared to those who received inactivated vaccines; however, no statistical significance was achieved (Figure 1b). Among patients who had received 3 doses of inactivated vaccines, the overall plasma neutralizing titer of moderate patients was higher than that of mild patients (Figure 1c).

**Figure 1.**
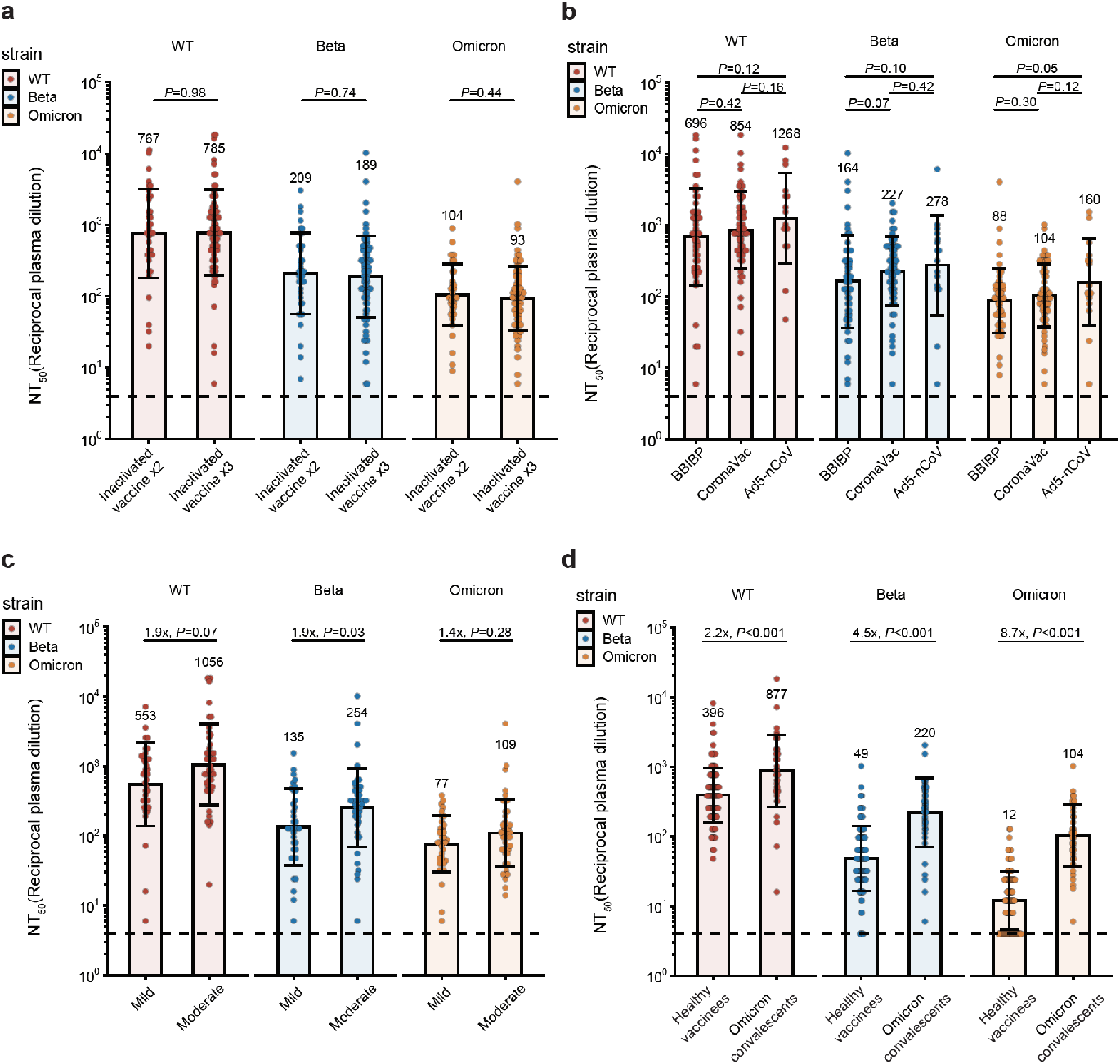
Humoral immune responses against WT and variants of SARS-CoV-2 among breakthrough infection convalescents. Plasma neutralization against authentic SARS-CoV-2 wildtype strain, Beta strain, and Omicron strain is displayed. The geometric mean titer (GMT), geometric standard deviation, and fold-changes of 50% neutralization titers (NT50) are labeled. Dashed lines show the limit of detection. Statistical significance was determined by a two-tailed Wilcoxon test. (a) NT50 from the breakthrough infection convalescents who were vaccinated with 2 (n=39) or 3 (n=80) doses of inactive vaccine. (b) NT50 from the breakthrough infection convalescents who were vaccinated with BBIBP-CorV (n=54), CoronaVac (n=65) or Ad5-nCoV (n=16) vaccine. (c) NT50 from the breakthrough infection convalescents with mild (n=36) or common (n=43) symptoms after 3 doses of inactive vaccine immunization. (d) NT50 from the breakthrough infection convalescents who were vaccinated with 3 doses of CoronaVac vaccine (n=42) or from the uninfected vaccinees who were vaccinated with 3 doses of CoronaVac vaccine (n=114).

We compared the Omicron convalescent patients who had received 3 doses of CoronaVac (n=42) with the healthy individuals who were also vaccinated with 3 doses of CoronaVac (n=114) regarding the neutralizing antibody titers against WT, Beta and Omicron BA.1. The healthy volunteers were selected to have a matched vaccination profile with the Omicron convalescents. For the 42 patients who had received 3 doses of CoronaVac, the median interval between receipt of the second dose and the third dose was 193.5 days (IQR 187.0-212.8); the median interval between receipt of the booster dose and infection was 55.5 days (IQR 37.8-75.0); the median interval between receipt of the booster dose and sampling was 95.5 days (IQR 80.5-116.75). For the healthy vaccinated cohort, the median interval between receipt of the second dose and the booster dose was 194.5 days (IQR 187.0-210.2); the median interval between receipt of the booster dose and sampling was 93.5 days (IQR 78.0-113.0). The geometric mean of the NT50 of Omicron convalescent patients was 2.2, 4.5 and 8.7 times that of the healthy vaccinated individuals when neutralizing WT, Beta and Omicron BA.1, respectively (Figure 1d). These observations suggest that infection with Omicron could greatly elevate plasma neutralizing titers, enabling a humoral immunity barrier against Omicron vairants.

### Risk and Protective factors of COVID-19

Ordered multi-class logistic regression model was constructed based on age, gender and number of vaccine doses (Figure 2). In all age groups of patients, advanced age is a risk factor for severe disease (OR 1.063, 95% CI 1.048-1.078, P<0.001). Neither gender nor inactivated vaccine was significantly correlated with disease severity. Among adult patients, age is an adverse factor for severe disease (OR 1.044, 95% CI 1.024-1.064, P<0.001), while 3 doses of inactivated vaccine is a protective factor (OR 0.227, 95% CI 0.065-0.787, P=0.020). However, for child patients, no significant correlation was observed between age/gender/vaccination and disease severity.

**Figure 2.**
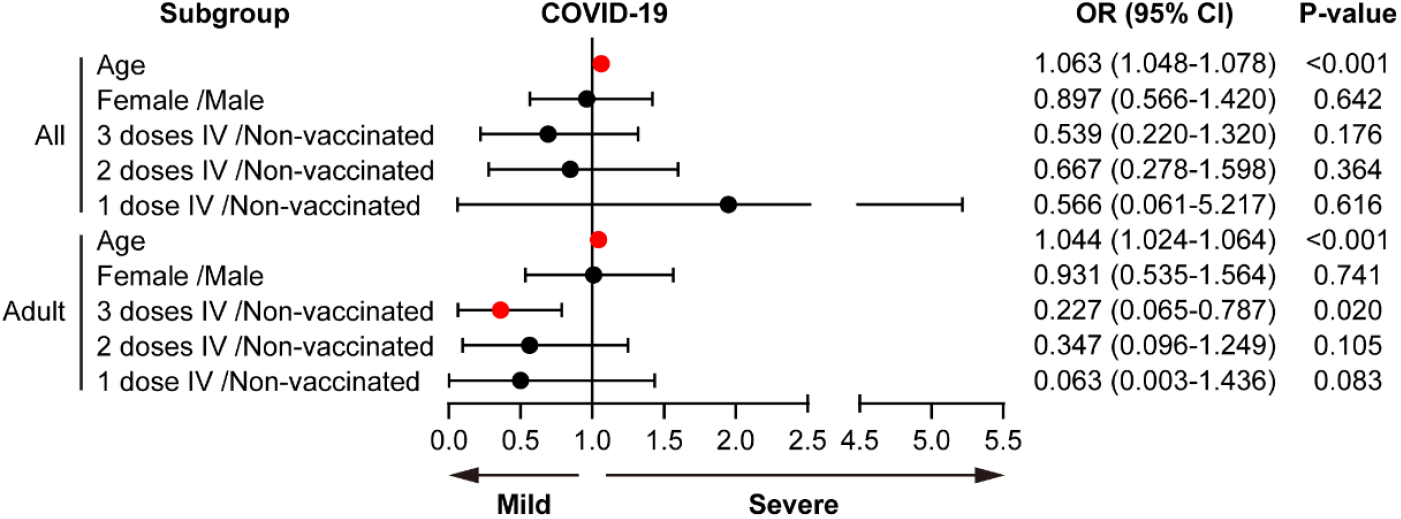
Risk and protective factors of COVID-19 disease severity according to the subgroup of Omicron-infected individuals.

Binary logistic regression suggests, that receipt of 3 doses of inactivated vaccine (as compared with no vaccination) is an independent protective factor against ICU admission for patients of all ages (OR 0.025, 95% CI 0.003-0.231, P=0.001). The same applies to adult patients alone (OR 0.023, 95% CI 0.002-0.214, P=0.001) (Figure S1). Receipt of 3 doses of inactivated vaccine is an independent protective factor against re-positive PCR for patients of all ages (OR 0.317, 95% CI 0.144-0.700, P=0.004) and adult patients (OR 0.240, 95% CI 0.098-0.587, P=0.002) (Figure S2). Receipt of 3 doses of inactivated vaccine is associated with shorter hospitalization and recovery (OR 0.415, 95% CI 0.201-0.853, P=0.017), even when adjusted for age and gender. The same also applies to adult patients alone (OR 0.233, 95% CI 0.091-0.596, P=0.002) (Figure S3).

## Discussion

This has been the first study to report on the protective effect of inactivated vaccines against severe disease caused by the Omicron variant. Particularly, findings from this study suggest that full vaccination (3 doses) of inactivated vaccine would substantially lower the risk of severe disease caused by Omicron in adult patients. No cases of severe COVID-19 were observed in fully vaccinated adult patients.

Previous studies reported a substantial decline of Omicron’s replication ability in lung tissues,^7^ and animal experiments found that viral RNA load is low in the lungs after infection, and no bronchitis or lung inflammation was observed.^8^ Such evidence suggests that Omicron causes less damage to the lungs and is less likely to result in severe pneumonia and respiratory distress, which may be the reason for a reduced proportion of severe and moderate cases. However, our analysis revealed that compared with unvaccinated patients, patients who have received 3 doses of inactivated vaccine before infection are substantially less likely to progress to ICU, have a lower risk of re-positive nucleic acid test and a shorter course of illness, suggesting that receipt of inactivated vaccine confers strong protection against severe disease.

Analysis of laboratory findings at the beginning of the convalescent phase indicated that, patients who had received a booster dose of inactivated vaccine tend to experience earlier inflammatory regression compared to unvaccinated patients, reflected in the lower systemic inflammatory index, C-reactive protein levels and relatively lower proportion of lymphocytes within a normal range. It has been reported that patients with severe COVID-19 are more prone to leukopenia and lymphopenia, and overly reduced lymphocyte percentage can be used as an indicator of disease severity.^9, 10^ However, based on overall immune characteristics of patients during the convalescence phase, we believe these laboratory findings point to lower degree of inflammation, suggesting that 3 doses of inactivated vaccine may shorten the course of illness by inducing resolution of inflammation. Receipt of booster dose is associated with an elevated proportion of activated Treg and Th1/Th2 ratio, suggesting that vaccination and infection may activate immune responses also by inducing spike-specific Th1 responses.^11^ This implies that even if mutations on Omicron spike protein affect T-cell epitopes, immune responses mediated by T cells or non-neutralizing antibodies can still provide protection.^12, 13^

Receipt of 3 doses of inactivated vaccine (CoronaVac) would induce a certain level of neutralizing antibodies against the Omicron variant. Omicron convalescent patients showed even more pronounced elevation in antibody titers regardless of the types of vaccine (inactivated or adenovirus-vectored) received prior to infection and the status of a booster vaccination.

Elevated titers of neutralizing antibodies in Omicron convalescent patients’ plasma indicate that previous infection can provide certain immune protection against SARS-CoV-2 reinfection. Although the previous infection does confer protection against reinfection, such protective effect is less pronounced in preventing reinfection by Omicron.^14^ This is also reflected in the observation that the increase in neutralizing titer (against Omicron) is relatively mild after Omicron infection (compared to previous variants), which is probably due to the fact that Omicron is less effective at antagonizing the host cell interferon response,^15^ leading to mild clinical symptoms.^16, 17^ These findings indicate weak immunogenicity of the Omicron variant, and reduced immune response makes reinfection possible.

The Tianjin outbreak driven by Omicron was centered around after-school training institutions for primary school students, leading to cluster infections. We analyzed the clinical characteristics of child patients who accounted for a large proportion of affected individuals. Most of the child patients had mild or no symptoms, only 12.3% with imaging findings of pneumonia, and none progressed to ICU. This could be attributed to weakened pathogenicity of Omicron^18^ or because most of the child patients (82.5%) had received 2 doses of inactivated vaccine. However, due to the limited sample size, resistance to the virus among different age groups of children cannot be inferred precisely. Yet, we found that age was a risk factor for severe COVID-19 disease. Up to 87.8% of child patients presented with merely mild symptoms or no symptoms, compared with 35.4% of adult patients. This indicates that people of older age are more likely to develop severe disease after infection, and strengthening the protection of middle-aged and senior populations would effectively reduce the burden of the pandemic on public health.

Of the 430 cases reported in this study, 85 experienced re-positive results on nucleic acid tests after two RT-PCR assays yielded a Ct value beyond 40. Currently, there is no agreement on the threshold of Ct value for positive results. For example, a Ct value ≥ 35 would be regarded as negative in the US, Canada and Japan, and ≥ 30 in Germany.^19^ Even when re-positive patients are defined by such standards, the re-positive rate among vaccinated patients remains low (Table S6). No secondary infection was caused by the re-positive cases and no recurrence or worsening of symptoms was observed, which is a major difference from reinfection cases. This indicates that discrete elevation in Ct value is a cross-sectional manifestation over the course of viral infection. Positive results on RT-PCR assay indicate the presence of nucleic acid fragments of the virus, but it does not necessarily suggest the presence of whole viruses or replication ability of the virus. Virus culture, whole-genome sequencing (WGS), and joint parallel detection of viral antigens or virus-specific nucleic acid sequences can be used to evaluate replication activity of the virus, and the detection of host antibodies produced against viral antigens in serum samples can infer the risk of continuous viral infection.

This study has several limitations. First, the sample of patients is small. Due to effective control efforts, the outbreak was soon contained and the number of cases was thus limited. We covered all cases in our analysis, including asymptomatic patients, to reflect the overall characteristics across all age groups. Second, WGS was performed in the first two cases to confirm that the causative agent was Omicron BA.1, and other cases were regarded as Omicron BA.1 infection based on epidemiological evidence. Tianjin reported the first domestic case of Omicron BA.2 within its municipal area on February 24, 2022, and the other 479 COVID-19 cases reported between February 24 and March 22, 2022 were not included in this study.

This study revealed epidemiological and clinical characteristics of the Tianjin outbreak driven by Omicron variant. Child patients tend to present with mild symptoms compared to adults. Full vaccination of the population can reduce the incidence of Omicron infections. Booster dose with inactivated vaccine can provide protection by inducing effective neutralizing antibodies against Omicron variant, reducing the risk of severe disease and ICU admission, and shortening the duration of illness.

## Data Availability

All data produced in the present study are available upon reasonable request to the authors

## Contributors

ZS conceived the study. XC, HZ and YX extracted and analyzed the clinical data. YH, YC, and YS collected the convalescent plasma samples. YC and YF provided the plasma of healthy vaccinated individuals. YC and WS analyzed the plasma neutralization titers. FW and SL performed the immune characteristics studies. WJ and XSX supervised the study. YC, XC, YX and ZS wrote the original draft with inputs from all authors. QG revised the manuscript.

## Data sharing

Although all data used in this analysis were anonymized, the individual level nature of the data used risks individuals being identified, or being able to self-identify, if the data are released publicly. Extracted data are available upon request.

## Acknowledgments

This work is funded by Changping Laboratory (CPL-1233) and the Emergency Key Program of Guangzhou Laboratory (EKPG21-30-3), sponsored by Ministry of Science and Technology of People’s Republic of China. We thank Sinovac Biotech for the assistance with authentic virus experiments. We thank the medical staff and laboratory staff of Tianjin Haihe Hospital and Tianjin First Central Hospital for their clinical work and laboratory tests. We thank all the volunteers involved in this study for providing critical information.

## Supplementary Appendix

**Table S1.**
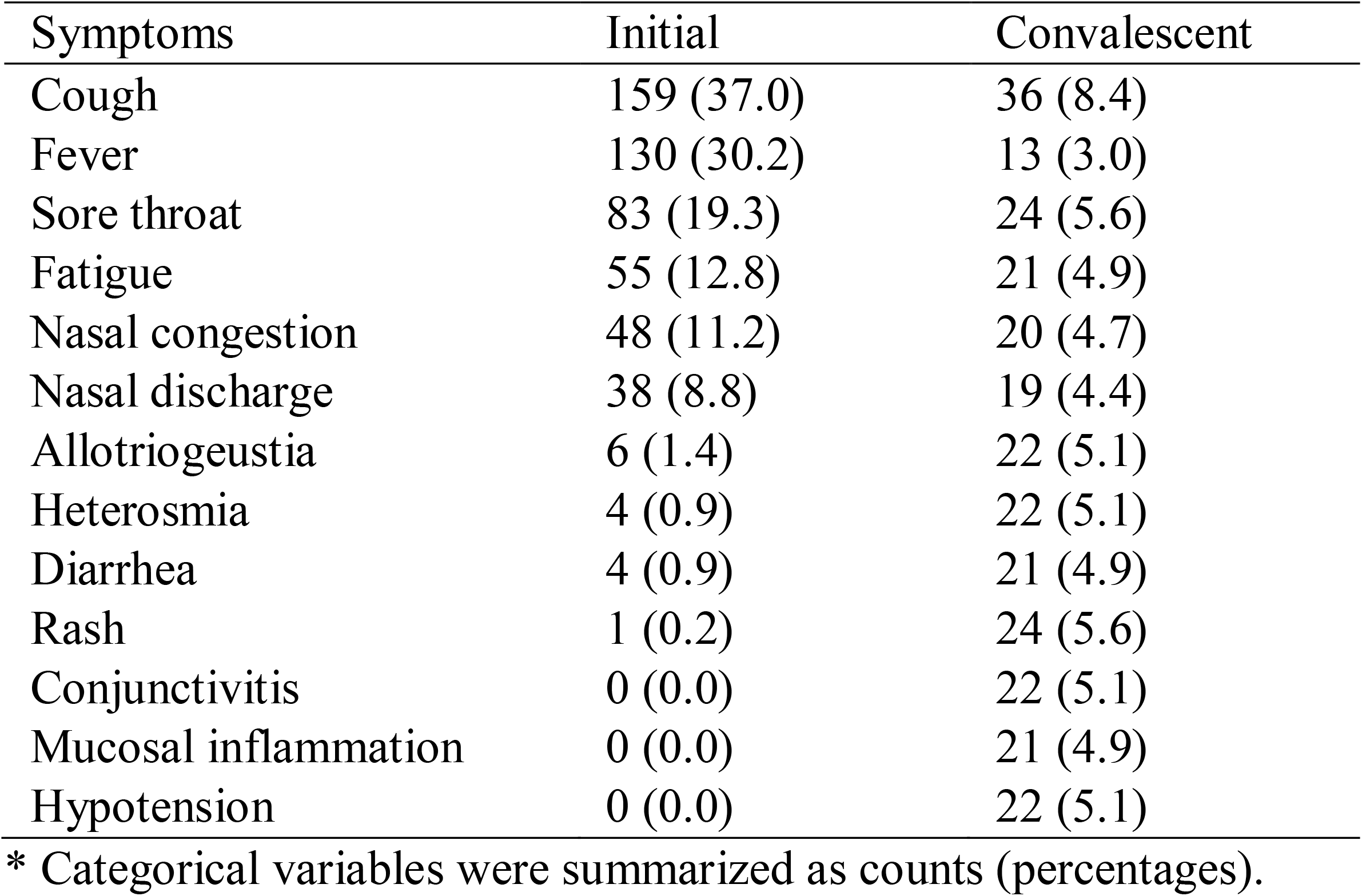
Initial symptoms and convalescent symptoms *.

**Table S2.** Comorbidities *.

| Disease |  |  |
| --- | --- | --- |
| Cardiovascular system | Hypertension | 73 (17.0) |
|  | Coronary artery disease | 18 (4.2) |
|  | Arrhythmia | 9 (2.1) |
|  | Other | 4 (0.9) |
| Digestive system | Abnormal liver function | 69 (16.0) |
|  | Fatty liver | 17 (4.0) |
|  | Gastrointestinal diseases | 12 (2.8) |
|  | Gallstone | 7 (1.6) |
|  | Other | 7 (1.6) |
| Endocrine system | Diabetes | 31 (7.2) |
|  | Electrolyte disorder | 20 (4.7) |
|  | Hyperlipidemia | 19 (4.4) |
|  | Thyroid dysfunction | 12 (2.8) |
|  | Other | 11 (2.6) |
| Respiratory system | Lung disease | 20 (4.7) |
|  | Bronchiectasia | 3 (0.7) |
| Nervous system | Cerebrovascular disease | 9 (2.1) |
|  | Viral encephalitis | 1 (0.2) |
|  | Malignant tumor of brain | 1 (0.2) |
| Genitourinary system | Chronic nephritis | 3 (0.7) |
|  | Renal malignancy | 1 (0.2) |
| Hematopathy | Anemia | 11 (2.6) |
|  | Leucopenia | 4 (0.9) |
|  | Thrombocytopenia | 3 (0.7) |
|  | Leukemia | 1 (0.2) |
| Dermatosis |  | 3 (0.7) |
| Other |  | 14 (3.3) |

\* Categorical variables were summarized as counts (percentages).

**Table S3.** Correlation between comorbidities and severity of COVID-19 diseases.

|  | Comorbidities<br>(N=172) | No comorbidities<br>(N=258) | P-value |
| --- | --- | --- | --- |
| Severity * |  |  | <0.001 |
| Asymptomatic | 1 (0.6) | 6 (2.3) |  |
| Mild | 50 (29.1) | 155 (60.1) |  |
| Moderate | 119 (69.2) | 97 (37.6) |  |
| Severe | 2 (1.2) | 0 (0.0) |  |
| Critical | 0 (0.0) | 0 (0.0) |  |

\* COVID-19 disease severity was defined according to WHO living guidance for clinical management of COVID-19. Categorical variables were summarized as counts (percentages).

**Table S4.** Correlation between PCR re-positive and convalescent symptoms.

|  | Convalescent symptoms<br>(N=60) | No convalescent symptoms<br>(N=370) | P-value |
| --- | --- | --- | --- |
| PCR re-positive * |  |  | 0.004 |
| Yes | 20 (33.3) | 65 (17.6) |  |
| No | 40 (66.7) | 305 (82.4) |  |

\* PCR, polymerase chain reaction for SARS-CoV-2. Categorical variables were summarized as counts (percentages).

**Table S5.** Correlation between hepatic and renal function and vaccination*.

|  | 3 Doses<br>(N=157) | 2 Doses<br>(N=178) | 1 Dose<br>(N=6) | Non<br>(N=38) | P-value |
| --- | --- | --- | --- | --- | --- |
| Glutamic pyruvic transaminase — IU/liter | 32.29 (20.32-67.33) | 18.91 (12.53-40.79) | 15.92 (12.06-28.17) | 16.94 (13.85-29.29) | <0.001 |
| Glutamic oxalacetic transaminase — IU/liter | 29.00 (23.14-37.88) | 26.24 (22.96-34.05) | 28.76 (27.16-31.73) | 31.72 (25.79-39.11) | 0.072 |
| Total bilirubin — µmol/liter | 12.33 (9.78-13.81) | 11.68 (9.61-14.00) | - | 10.62 (8.47-13.61) | 0.322 |
| Gamma-glutamyl transpeptidase — IU/liter | 25.16 (17.33-54.26) | 25.80 (15.14-51.63) | - | 18.01 (14.47-38.44) | 0.499 |
| Alkaline phosphatase — IU/liter | 68.12 (56.43-81.84) | 74.89 (61.82-94.30) | - | 79.67 (54.00-124.76) | 0.133 |
| Albumin — g/liter | 42.73 (40.91-45.46) | 41.43 (38.62-44.72) | - | 36.12 (31.97-43.31) | 0.008 |
| Creatinine — µmol/liter | 61.33 (50.86-71.81) | 46.66 (38.31-58.80) | 38.11 (37.61-59.12) | 43.52 (32.30-58.82) | <0.001 |
| Urea nitrogen — mmol/liter | 3.97 (3.35-4.69) | 3.94 (3.10-4.68) | 5.19 (4.06-5.65) | 4.53 (3.48-5.87) | 0.011 |
| UREA/CREA † | 0.07 (0.06-0.08) | 0.08 (0.06-0.10) | 0.12 (0.10-0.13) | 0.11 (0.07-0.14) | <0.001 |

\* Data were missing for the measurement of glutamic pyruvic transaminase, glutamic oxalacetic transaminase, creatinine, urea nitrogen and UREA/CREA in 2 patients (0.5%), for total bilirubin, glutamyl transpeptidase, alkaline phosphatase and albumin in 322 patients (74.9%). Continuous variables were shown in median (interquartile ranges).
† UREA/CREA was defined as the ratio of plasma urea nitrogen to plasma creatinine.

**Table S6.** Correlation between PCR re-positive and vaccination *.

|  | Vaccinated<br>(N=392) | Non-vaccinated<br>(N=38) | P-value | Adult<br>(N=316) | Child<br>(N=114) | P-value |
| --- | --- | --- | --- | --- | --- | --- |
| PCR Ct<35 |  |  | 0.005 |  |  | 0.041 |
| Yes | 63 (16.1) | 13 (34.2) |  | 63 (19.9) | 13 (11.4) |  |
| No | 329 (83.9) | 25 (65.8) |  | 253 (80.1) | 101 (88.6) |  |
| PCR Ct<30 |  |  | 0.001 |  |  | 0.047 |
| Yes | 16 (4.1) | 7 (18.4) |  | 21 (6.6) | 2 (1.8) |  |
| No | 376 (95.9) | 31 (81.6) |  | 295 (93.4) | 112 (98.2) |  |

\* PCR, polymerase chain reaction for SARS-CoV-2. Categorical variables were summarized as counts (percentages).

**Figure S1.**
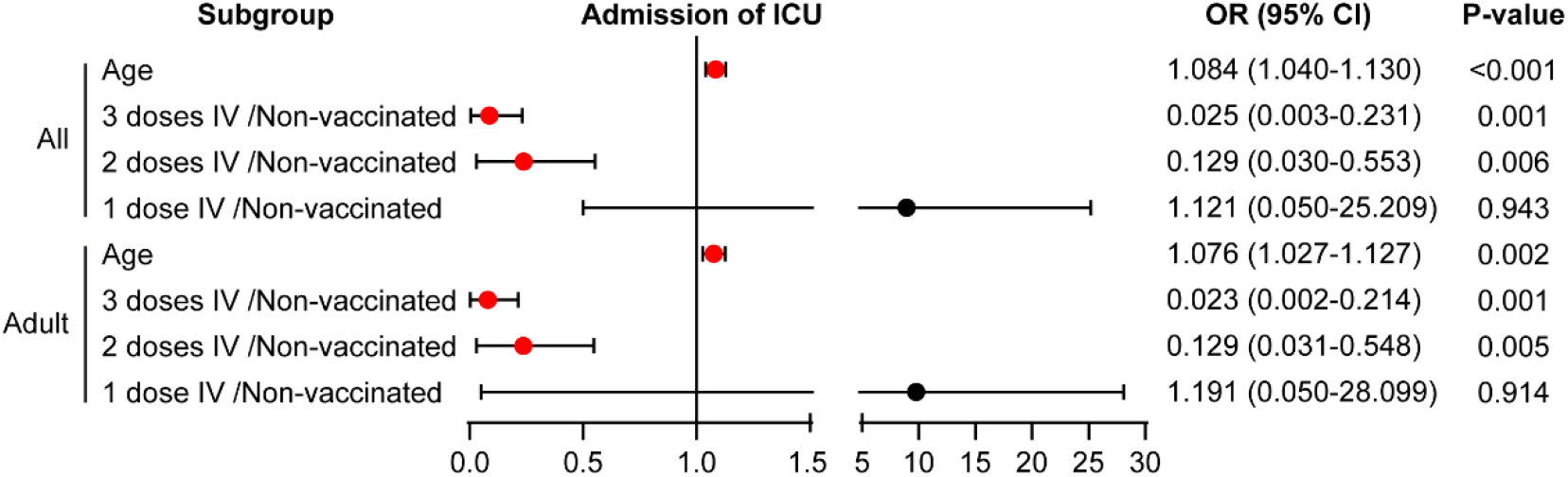
ICU admission according to subgroup in infected individuals by binary logistic regression.

**Figure S2.**
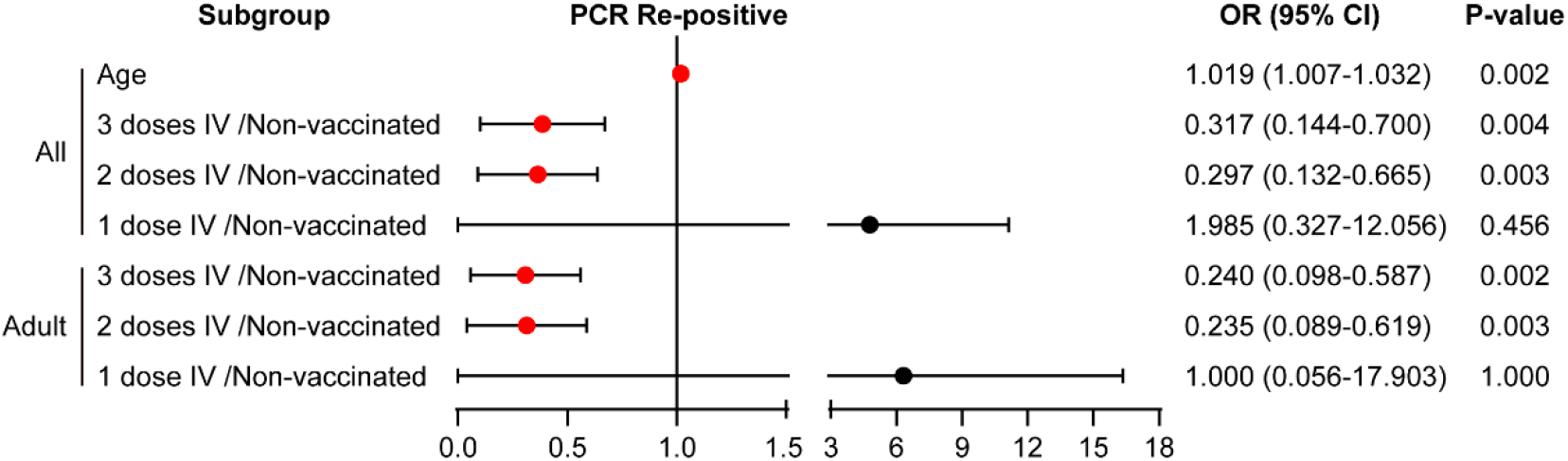
PCR re-positive status according to subgroups in infected individuals by binary logistic regression.

**Figure S3.**
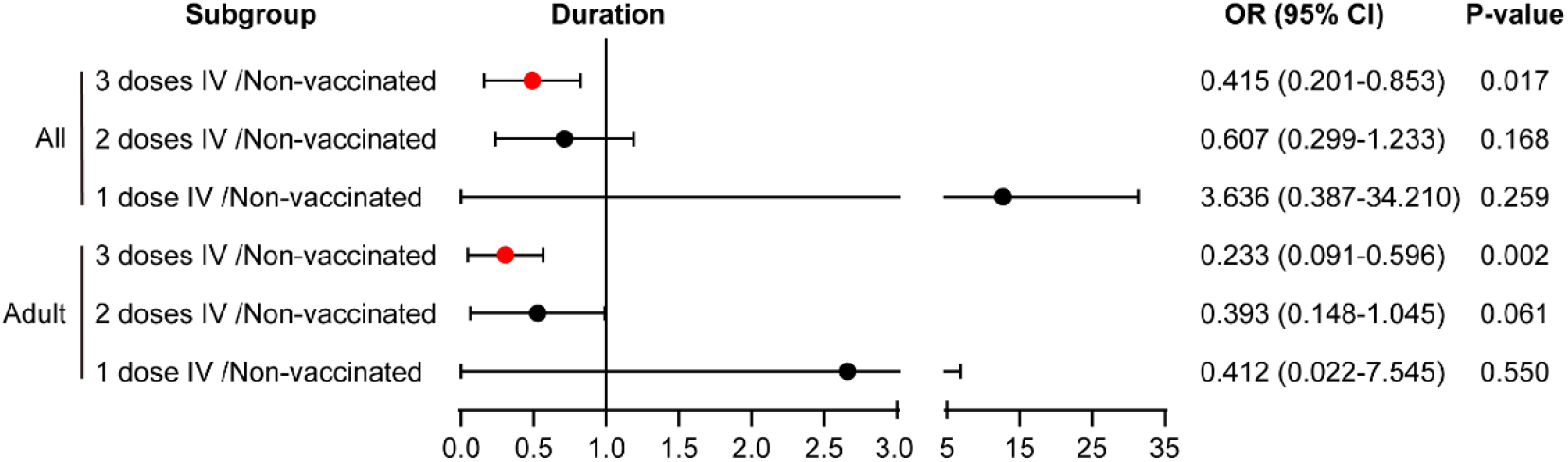
Hospital stay and rehabilitation duration according to subgroup in infected individuals by binary logistic regression.

